# Using Convergent Sequential Design for Rapid Complex Case Study Descriptions: Example of Public Health Briefings During the Onset of the COVID-19 Pandemic

**DOI:** 10.1101/2020.11.11.20229393

**Authors:** Cheryl N. Poth, Okan Bulut, Alexandra M. Aquilina, Simon J. G. Otto

## Abstract

Conceptualizing the public health response to the COVID-19 pandemic response as a complex adaptive system is useful to study its key features of emergence, interdependency, and adaptation yet practical guidance for mixed methods researchers remains limited. This study contributes an illustrative example and discussion for guiding how a mixed methods convergent sequential research design, informed by complexity theory and drawing upon open-access datasets, can rapidly generate complex case study descriptions. This article serves as an essential reference for identifying points of integration within a sequential convergent design using text mining to manage large data volumes and studying complex phenomena using a complexity-informed case study-mixed methods approach to generate novel public health insights.

---

This study contributes an illustrative example and discussion for guiding how a mixed methods convergent sequential research design, informed by complexity theory and drawing upon open-access datasets, can rapidly generate complex case study descriptions. The case studied in this article draws upon open-access datasets to examine how public health briefings can be used to build the necessary credibility and trust for effective public health communications during the unprecedented and rapidly changing conditions surrounding the onset of the COVID-19 pandemic in Alberta (Canada). In the convergent sequential design, qualitative and quantitative findings are integrated in the follow-up phase for the purpose of generating a more complete understanding of the initial phase findings. Specifically, this article focuses on the procedures involved in the follow-up phase generating the meta-inferences from the cross analysis of the integrated findings at six key fluctuation periods identified in the initial quantitative phase. Full details of the initial quantitative phase and discussion of the integrated findings are provided elsewhere (see Bulut & Poth, 2020; Poth, Bulut, Otto, & Aquilina, 2020). We conclude this article with discussing how attending to three key concepts of complex adaptive systems— emergence, interdependency, and adaptation—informed the generation of the complex case description. To begin, we present some background information on the public health crisis surrounding the COVID-19 global pandemic to explain the conceptualization of the public health response as a complex adaptive system (CAS) and the overall aim of the intrinsic case study of the onset of the global pandemic in Alberta (Canada) to inform effective public health communications. Then we explain the usefulness of complexity theory for studying a CAS and the application of a case study-mixed methods design with a nested convergent sequential approach for generating complex case descriptions quickly.

## The Global Public Health Crisis Surrounding The COVID-19 Pandemic Response as a Complex Adaptive System

The public health crisis surrounding COVID-19—an infectious disease caused by a novel coronavirus named severe acute respiratory syndrome (SARS) coronavirus 2—and the declaration of a pandemic by the World Health Organization (WHO) on March 11, 2020, disrupted and changed in nonlinear and unpredictable ways how people around the world live their lives and interact with others. Phenomena that defy simplistic analyses of cause and effect and that have the capacity to adapt to contextual changes are known as a CAS (Weaver, 1948). Conceptualizing the public health response to the COVID-19 pandemic response as a CAS is useful to guide its study because it is influencing and being influenced by a large number of interacting and interrelated contexts for which there is no central control and such as response requires ongoing adaptation to changing circumstances. A key component of the initial public health response to the COVID-19 global pandemic involved the use of regular publicly accessible media briefings led by public health officials representing regional, national and international health authorities and government bodies. These media briefings became a key source of information about emerging understandings about the virus transmission rates and methods as well as changes to risk factors and preventive measures that needed to be conveyed in a timely and accurate manner within the rapidly evolving local, national, and international circumstances.

Studying the public health response to this unprecedented global pandemic as a CAS requires attending to the key concepts of a CAS in the following three ways: First, that *emergence* results from the ability of a CAS to self-organize and be affected by other influences means that communications will necessarily convey new understandings about the virus as they become available. Second, that *interdependency* results from systems being interconnected and parts entwined with other parts of society means that the public health communications will necessarily involve information about various sectors as well as being influenced by and influencing to various sectors (e.g., industries, restaurants, continuing care facilities, hospitals, and others). Third, that *adaptation* resulting from living systems surviving and thriving within their constantly changing environments means that the public health communications will necessarily need to reflect up-to-date information that is both accurate and timely. Therefore, the study of a CAS requires appropriate designs and data sources for measuring their nonlinear trajectories and assessing the patterns emerging within the systems that are continuously self-forming and interacting with each other. Generating in-depth descriptions and insights about the nature of the patterns of communication and how the patterns changed and evolved over time was the focus of the current empirical study with the overall aim of informing effective communication practices during the onset of a global pandemic.

## The Usefulness of Complexity Theory for Studying a CAS Using a Case Study-Mixed Methods Design

Complexity theory is an umbrella term for the study of complexity and complex systems. A CAS is a special case of complex systems where the whole is more complex than its parts (Holland, 1999). The study of CAS requires new design approaches because it is no longer possible to study only the parts of the system and the system itself in isolation. Instead it is necessary to study the interconnections including the possible influences to and results from the interactions within the system and surrounding systems (Poth & Bullock, 2020). Case study and mixed methods, alone as well as in combination, are well established research approaches known to require extensive time to complete (Creswell & Plano Clark, 2018; Guetterman & Fetters, 2018). A case study involves developing an in-depth description of a case within its real-life bounded system often defined by place, time, and people (Creswell & Poth, 2017). Case studies draw upon diverse data sources relevant to the case studied and can involve the integration of both qualitative and quantitative research in the case description (Yin, 2017). In the widely utilized typology advanced by Creswell and Plano Clark (2018), mixed methods case studies are included as one of four complex designs and described as particularly useful for studying complex systems. Our study is guided by the innovative distinction of the designation case study-mixed methods (CS-MM) design (Gueterman & Fetters, 2018) where the parent case study approach is characterized by an intrinsic purpose, a holistic case description, and a nested mixed methods design as a means to address the time constraints associated with data collection, analysis, and interpretations.

Our study design addressed the feasibility challenges we experienced as researchers to keep pace with the rapidly evolving context of the global pandemic. With the overall aim of generating a comprehensive understanding of the patterns of effective, rapid public health communications, we use a CS-MM to guide our development of a complex case description through gathering and integrating diverse sources of data drawing upon open-access datasets and text mining techniques. Recently, the availability of existing big data stored in the form of text has motivated researchers to apply modern text mining techniques (e.g., sentiment analysis, topic modeling, and text classification) to organize, analyze, and gain insights from textual data. The primary goal of text mining with big data is to analyze information to discover patterns (Aggarwal & Zhai, 2012). The further use of data and text mining techniques has been identified as an emerging area for methodological innovations in mixed methods research (Creswell & Plano Clark, 2018; Poth, 2018).

## Application of a Complexity-Informed CS-MMR Design with a Nested Convergent Sequential Approach for Generating Complex Case Descriptions Quickly

A convergent sequential design represents a departure from the three core design typologies articulated by Creswell and Plano Clark (2018). Common to both the convergent sequential design and the explanatory sequential design is an initial quantitative phase. The designs are distinguishable by the types of data and mixing purpose for the follow-up phase; whereas the explanatory sequential design traditionally uses qualitative results to explain the initial quantitative findings, the convergent sequential design uses the integration of quantitative and qualitative results to generate more complete understandings of the initial quantitative findings. The second phase of the convergent sequential design has a similar mixing purpose to the convergent design that is another core design typology described by Creswell and Plano Clark (2018).

The sequential aspect of the convergent sequential design allows researchers to find or choose a subsample from the larger quantitative results in the initial phase. Researchers have shown a great deal of interest in text mining techniques to process large volumes of data more quickly, extract hidden information, and use the knowledge in decision making (Aggarwal, 2015; Aggarwal & Zhai, 2012). We are not alone in our application of text mining to study the global pandemic. Recently researchers around the world have employed sentiment analysis to analyze sentiments and opinions that people shared about the COVID-19 pandemic through social media posts (e.g., Barkur et al., 2020; Medford et al., 2020; Samuel et al., 2020; Zhou et al., 2020). With the rapid growth of textual data in today’s digital world, text mining is likely to remain a crucial technique that social scientists will employ to better understand complex societal problems yet it remains an under-utilized approach within mixed methods research designs. In this study, the two sequential design phases realized the overall objective of generating a complex case description quickly; the first phase used text mining to identify key fluctuations in sentiment messaging and word counts across the case and the second phase involved the cross analysis of the integrated findings at each of the key fluctuation periods. The case description is guided by attending to the emergence, interdependencies, and adaptations of topic areas and message contents related to key areas of effective public health communications and risk assessments necessary to convince the public to follow recommendations during a pandemic (CDC, 2018; Tumpey et al., 2018). Through the use of a convergent sequential approach nested within a CS-MM design, this study begins to address the methodological and practical gaps by generating empirically based meta-inferences from open-access datasets.

## Method

In the convergent sequential mixed methods research design, qualitative and quantitative findings are integrated in the follow-up phase for the purpose of generating a more complete understanding of the initial phase’s findings. In this study, convergence in the follow-up phase was assessed across six key fluctuation periods identified in the initial phase to generate the meta-inferences informing the complex case description (see Table 1).

**Table 1.**
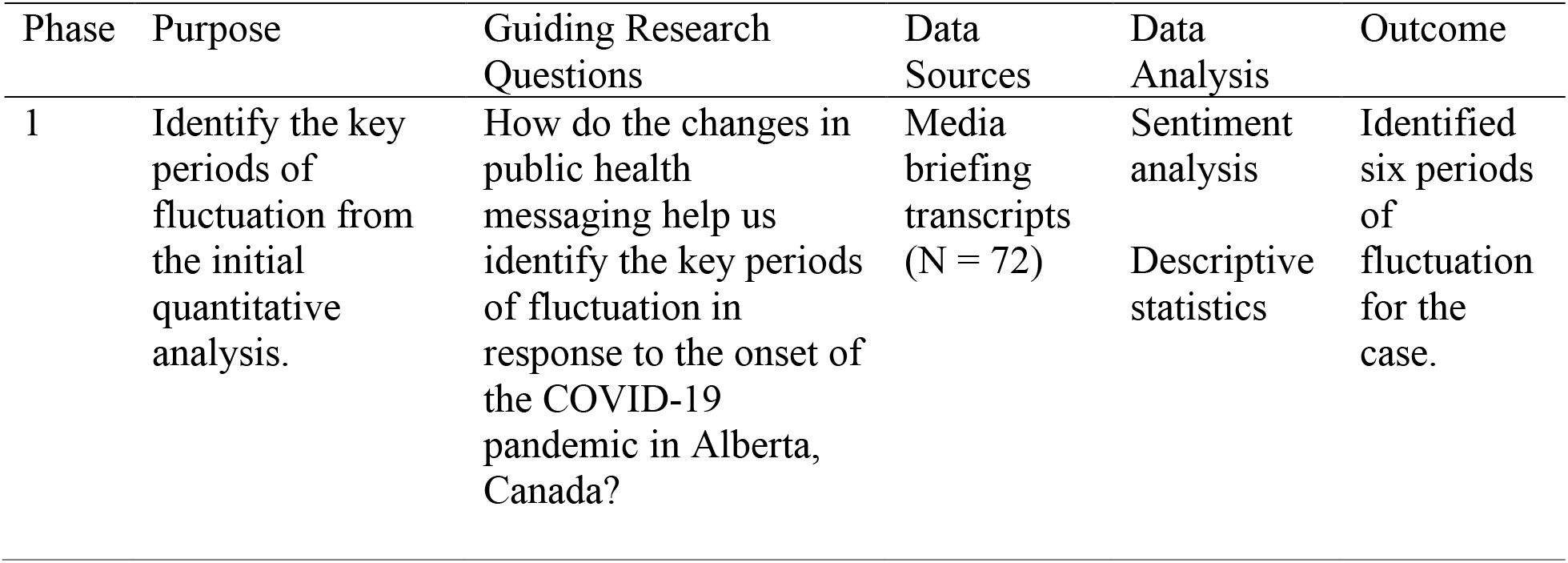
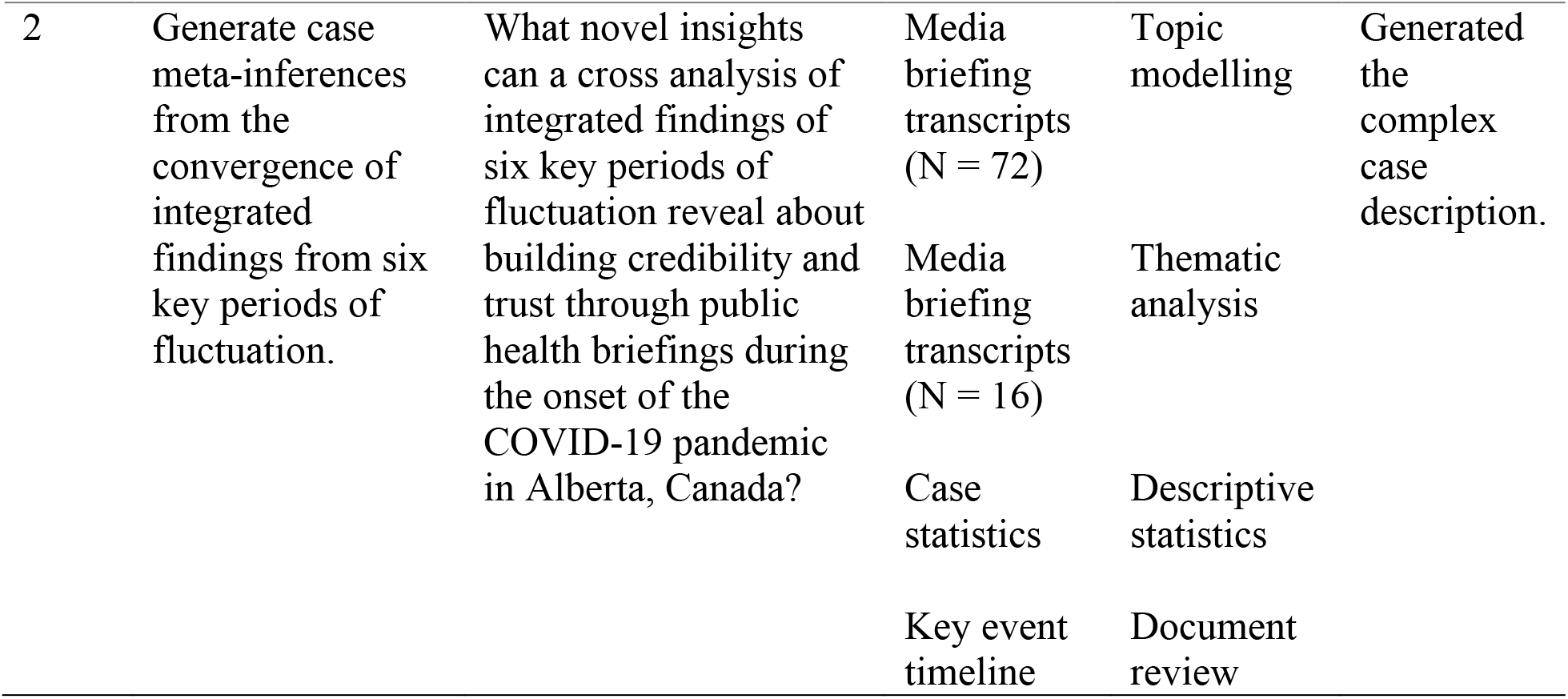
Purposes and Guiding Research Questions for the Two Phases of the Design.

| Phase | Purpose | Guiding Research Questions | Data Sources | Data Analysis | Outcome |
| --- | --- | --- | --- | --- | --- |
| 1 | Identify the key periods of fluctuation from the initial quantitative analysis. | How do the changes in public health messaging help us identify the key periods of fluctuation in response to the onset of the COVID-19 pandemic in Alberta, Canada? | Media briefing transcripts (N = 72) | Sentiment analysis<br>Descriptive statistics | Identified six periods of fluctuation for the case. |
| 2 | Generate case meta-inferences from the convergence of integrated findings from six key periods of fluctuation. | What novel insights can a cross analysis of integrated findings of six key periods of fluctuation reveal about building credibility and trust through public health briefings during the onset of the COVID-19 pandemic in Alberta, Canada? | Media briefing transcripts (N = 72) | Topic modelling | Generated the complex case description. |
|  |  |  | Media briefing transcripts (N = 16) | Thematic analysis |  |
|  |  |  | Case statistics | Descriptive statistics |  |
|  |  |  | Key event timeline | Document review |  |

### Selection and Bounding of the Intrinsic Case Studied

Alberta, a Western Canadian province offers an intrinsic context in which to study a localized response to the pandemic situated within a country with long-established democratic rights and values, freedom of the press, credible statistical sources, and a publicly-funded healthcare system. With a population of more than four million, Alberta represents approximately 11% of Canada’s total population with a smaller, on average, proportion of residents over the age of 65 (13% compared with 17.5% nationally) (Statistics Canada, 2019). In 2019, the *Economist* ranked Canada the seventh most democratic nation in its Democracy Index, tied with Denmark and ahead of all other nations in the Americas (Economist Intelligence Unit, 2019). Its mass media communications have a high level of media freedom evidenced by its ranking as sixteenth of 180 countries in the 2020 World Press Freedom Index (Reporters Without Borders, 2020). Formed in 1971, Statistics Canada is globally recognized as a producer of credible statistics for all the provinces as well as for the federal government; it regularly releases open-access datasets and provides statistical capacity-building around the world (Government of Canada, 2020a). Since 1962, Canada’s publicly funded healthcare system has covered all essential basic needs delivered through the ten provincial and three territorial systems, and is governed by the Canada Health Act adopted in 1984 (Government of Canada, 2020b).

The federal and provincial structures in the area of public health create unique opportunities for coordination; the Public Health Agency of Canada was established in 2004 amidst the coronavirus outbreak of SARS-CoV-1 and provides oversight at the national level. Dr. Teresa Tam has served since 2017 in the Chief Public Health Officer of Canada role as the lead health professional and primary spokesperson on public health related matters for the government of Canada (“Chief Medical Officer of Canada”, 2020). The vast majority of Canadians (88%) rated the national public health response to COVID-19 in August 2020 as “good” (Devlin & Connaughton, 2020) and cases and infection rates are considered by external assessments to be lower than many global counterparts (Bejan & Nicolova, 2020). A recent survey found Canadians have a high level of trust in public health officials on COVID-19 (Carleton, 2020)

When considering death rate as the single most fair and reliable statistic in ascertaining how hard hit any area has been hit by COVID-19, Alberta’s death rate at 58.6 per million is much lower when compared with other provinces (as of September 21, 2020): Quebec at 648 per million and Ontario at 194 per million (tableau public, September 15, 2020). The Chief Medical Officer of Health in Alberta serves as the primary spokesperson representing the Office of the Chief Medical Officer of Health and provides public health expertise to support health surveillance, population health and disease control initiatives on issues of public health importance to the province (Government of Alberta, 2020a). Dr. Deena Hinshaw was appointed to the role in 2019 and has been widely recognized as effective in her public health communications during the pandemic: “the wise and empathetic chief medical officer of health Dr. Deena Hinshaw deserves credit for helping to keep Alberta’s death rate so low while keeping the economy so open.” (Staples, 2020d).

We define the current case boundaries as the province of Alberta (location), March 5 to July 5, 2020, (onset), and Dr. Deena Hinshaw and her office (producers and communicators of the media briefings). The publicly accessible briefing transcripts provide a proxy for the communications from public health officials to members of the Alberta public. The onset date is defined by the first day the briefings were delivered on a regular (almost daily) basis until July 5, 2020.

### Data Source Selection and Collection

The study drew upon three sources of data: media briefing transcripts, case statistics, and a key event timeline (see Figure 1). This study did not require ethics approval because we used existing research datasets available through the public domain with no new data collected. To examine the public health communications hosted by Dr. Deena Hinshaw, media briefing transcripts were downloaded in Word format from the government of Alberta’s COVID-19 pandemic website (https://www.alberta.ca/covid). To examine the change in number of daily new Alberta confirmed cases of COVID-19, case statistics were downloaded and aggregated in excel spreadsheet format from the government of Alberta’s interactive COVID-19 data application website (https://www.alberta.ca/stats/covid-19-alberta-statistics.htm). To contextualize the public health messaging within the rapidly evolving and embedded local, national and international contexts surrounding the progression of the COVID-19 pandemic in Alberta, a timeline of key events was created from online news searches. Two researchers searched key dates and articles and entered them into a timeline table drawing upon new sources that were local (e.g., folio, Edmonton Journal), national (e.g., Global News, Canadian Medical Association Journal News, Canadian National Broadcasting Corporation (CBC), Canadian Healthcare Network), and international (e.g., British Broadcasting Corporation (BBC), Cable News Network (CNN), and World Health Organization WHO).

**Figure 1.**
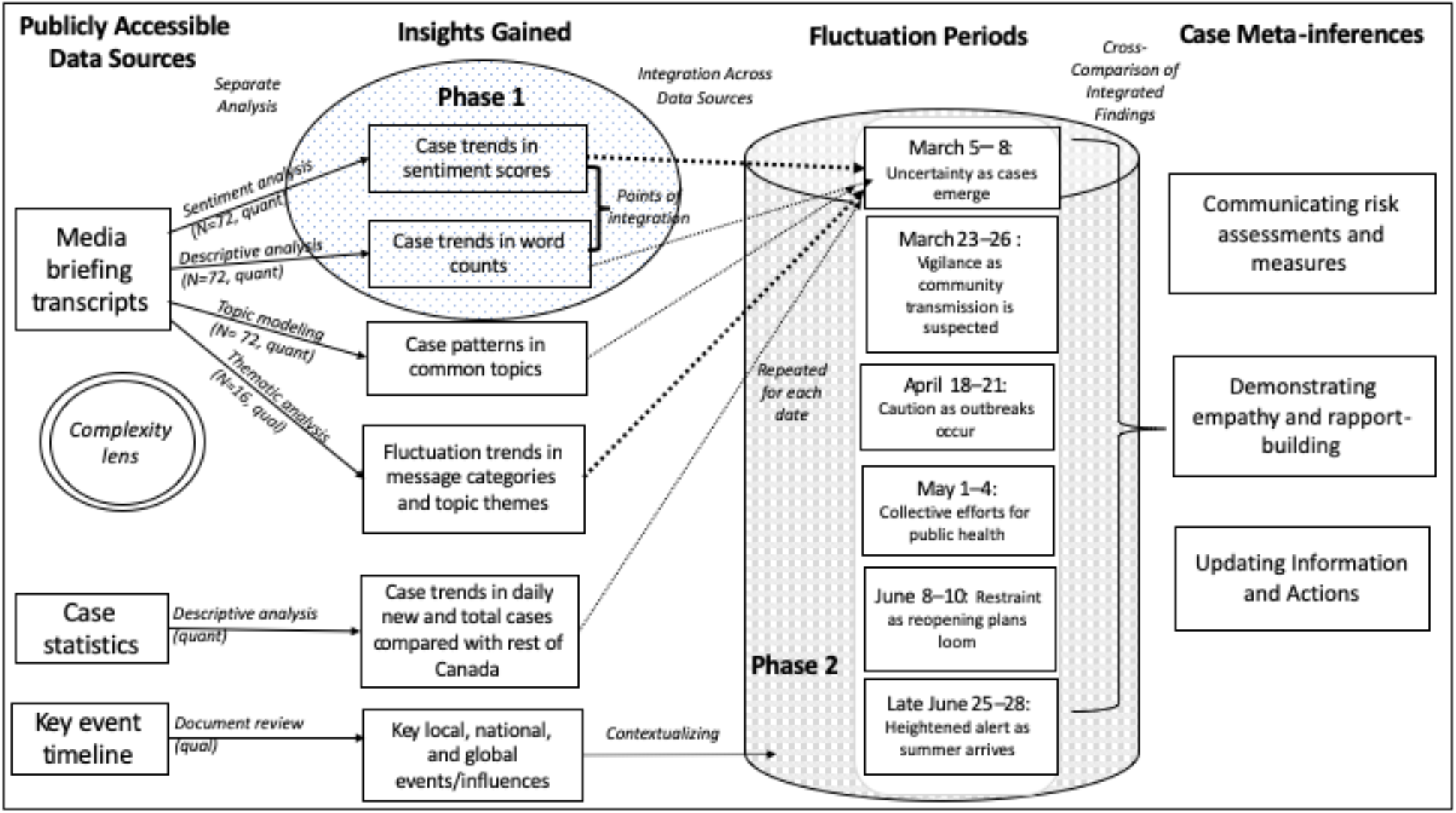
Design and Procedural Representation.

### Data Analysis and Integration Procedures

The first phase involved conducting sentiment and descriptive analysis of media briefing transcripts for the purpose of identifying key fluctuation periods across the case on which to integrate in the second phase. Sentiment analysis refers to the use of text mining techniques to identify positive and negative sentiments at the word, sentence, and document levels (Yi et al., 2003). For this study, we used the sentiment scores from a previous study in which sentiment analysis was applied to extract sentiment scores from the textual data in the media briefings (for a full description, see Bulut & Poth, 2020). In brief, word tokenization in the tidytext (Silge & Robinson, 2016) and tokenizer (Mullen et al., 2018) packages in R (R Core Team, 2020) were applied to sentences in the media briefings to split them into individual words. After filtering out stop words (e.g., the, a, that, on), the remaining words were merged with the Bing lexicon (Hu & Liu, 2004) in order to categorize the words as either positive or negative. For each media briefing, the sentiment score was calculated as the difference between the number of positive and negative words. Descriptive analysis involved the use of word counts in the media briefings. The goal of descriptive analysis was to identify the media briefings in which Dr. Hinshaw’s briefing was either very short or very long. Purposefully we defined the fluctuations to include four days to balance feasibility of analysis with the realities that case counts and news can be delayed by a day or two.

The second phase of the design involved conducting a cross analysis of the integrated findings at each of the key fluctuation periods identified in the first phase to generate the meta-inferences informing the complex case description. We used the qualitative dominant crossover mixed analysis (Onwuegbuzie & Hitchcock, 2015) to generate the integrated findings at each of the six key fluctuation periods across the case involving findings from four data sources: qualitative themes from the media briefing transcripts, key timeline events from the online news searches, key topics and associated terms from the quantitative topical modelling, and new case statistics from the quantitative descriptive analysis.

To begin, two of the authors independently conducted the qualitative analysis of the subset of media briefings organized chronologically by fluctuation period. This work was guided by a three stage coding cycle (Saldaña, 2015). In the first stage, we read all the transcripts delivered during key fluctuation periods (N = 16) to familiarize ourselves with the data, and then we undertook a line-by-line initial coding approach of the briefings (Charmaz, 2014). This open-ended coding method allowed us to remain aware of the possibilities emerging from the data and to develop a preliminary code list which was then reviewed by the authors, revised, and organized by five themes and 11 sub-themes (Miles, Huberman, & Saldaña, 2018). The codes were then applied to the subsequent transcripts for key fluctuation periods. A document analysis of the key events during each key fluctuation at local, national, and international levels were noted. Topic modeling was used to generate insights from the clustering of similar word groups and expressions that characterize the media briefings across the months using the topicmodels package (Grün & Hornik, 2011) in R (R Core Team, 2020). Topic modeling is an exploratory text mining technique to discover topics covered in a set of documents by grouping similar words and expressions (Ramage et al., 2009; Wallach, 2006). New case statistics was analyzed using R (R Core Team, 2020) to identify important daily fluctuations (e.g., spikes in new cases and sharp declines) in new confirmed cases of COVID-19. New case counts were chosen over the total number of cases because that enabled the detection of a large daily fluctuations in the number of confirmed cases, was more easily comparable to the Canadian statistics, and was deemed more accurate.

Integration was undertaken at each of the six key fluctuation periods using the qualitative themes as the organizational framework on which to converge the quantitative findings from topic modelling and case statistics to generate a more complete description of the key fluctuations; both were guided by a qualitative crossover mixed analysis approach (Onwuegbuzie & Hitchcock, 2015). The application of the concepts of emergence of new understandings, interdependency of the local, national, and international contexts, and adaptation of new measures and protocols meant that the processes involved in integration were iterative and spiral-like as the integrated findings were represented narratively in the case description. In so doing, we embraced the iterative flexibility described by Heinrich and colleagues (2016) and spiral metaphor proposed by Schoonenboom (2019) to respond to contextualizing opportunities for the key timeline events as connections were made between the narrative data to bring new meaning to the numerical data.

### Team Approach

Throughout the study, the interdisciplinary team of four researchers met to bring together their diverse expertise to move the research forward and in, so doing, embodied the concept of team integration described by Fetters and Molina-Azorin as one which “involves leveraging personal and professional background experiences that lead one to consider, and hold valuable, qualitative, quantitative, and mixed methods procedures for making sense of the world” (2017, p. 296). The first author brought extensive experience in applying complexity science to mixed methods as well as qualitative case research experiences to lead the overall design, integration and methodological purpose. The second author brought extensive quantitative and qualitative experiences to lead the first phase and contribute to implementation and writing. The third author was a doctoral student who was actively acquiring diverse experiences across diverse research methodologies and contributed to the data collection analysis and integration. The fourth author brought experience in emergency outbreak management and public health expertise. Together the team generated case meta-inferences that would otherwise have been inaccessible by the researchers working alone.

### Phase 1: Identification of Key Fluctuations Periods from Initial Quantitative Findings

The initial phase findings identified six key fluctuations periods to serve as the points of integration in the follow-up phase. Figure 2 and 3 presents the data trends in sentiment analysis word counts of the media briefings respectively. Table 2 summarizes the sentiment scores and descriptive results to justify the selection of the key fluctuation periods (see also Bulut & Poth, 2020 for full results description).

**Table 2.** Identification of Key Fluctuation Periods.

| Periods involved in key fluctuations | Sentiment Score Average (Range) | Word Count Average (Range) | Rationale for choice |
| --- | --- | --- | --- |
| Case (4 months) | -12.2<br>(-48-17) | 388<br>(132-729) | Case focus on onset of COVID-19 pandemic in Alberta, Canada |
| March 5-8 | -16.3<br>(-25--7) | **241**<br>(137-309) | Negative sentiment and lowest average word count |
| March 23-26 | -16.5<br>(-48-1) | 463<br>(366-593) | Most negative individual sentiment score, greatest change in sentiment score and second highest average word count |
| April 18-21 | -25.3<br>(-37--13) | 336<br>(132-522) | Negative sentiment, lowest individual word count, and greatest increase in word count |
| May 1-4 | -41.5<br>(-47--36) | **628**<br>(527-729) | Most negative average sentiment score, highest average word count, and highest individual word count |
| June 8-11 | -4<br>(-25-17) | 399<br>(398-401) | Most positive individual sentiment score, most positive sentiment average score and close to the case average word count |
| June 25-30 | -14.5<br>(-28--1) | 327<br>(297-356) | Negative sentiment and close to the case sentiment score average |

*Note.* **Bolded** numbers represent extremes.

**Figure 2.**
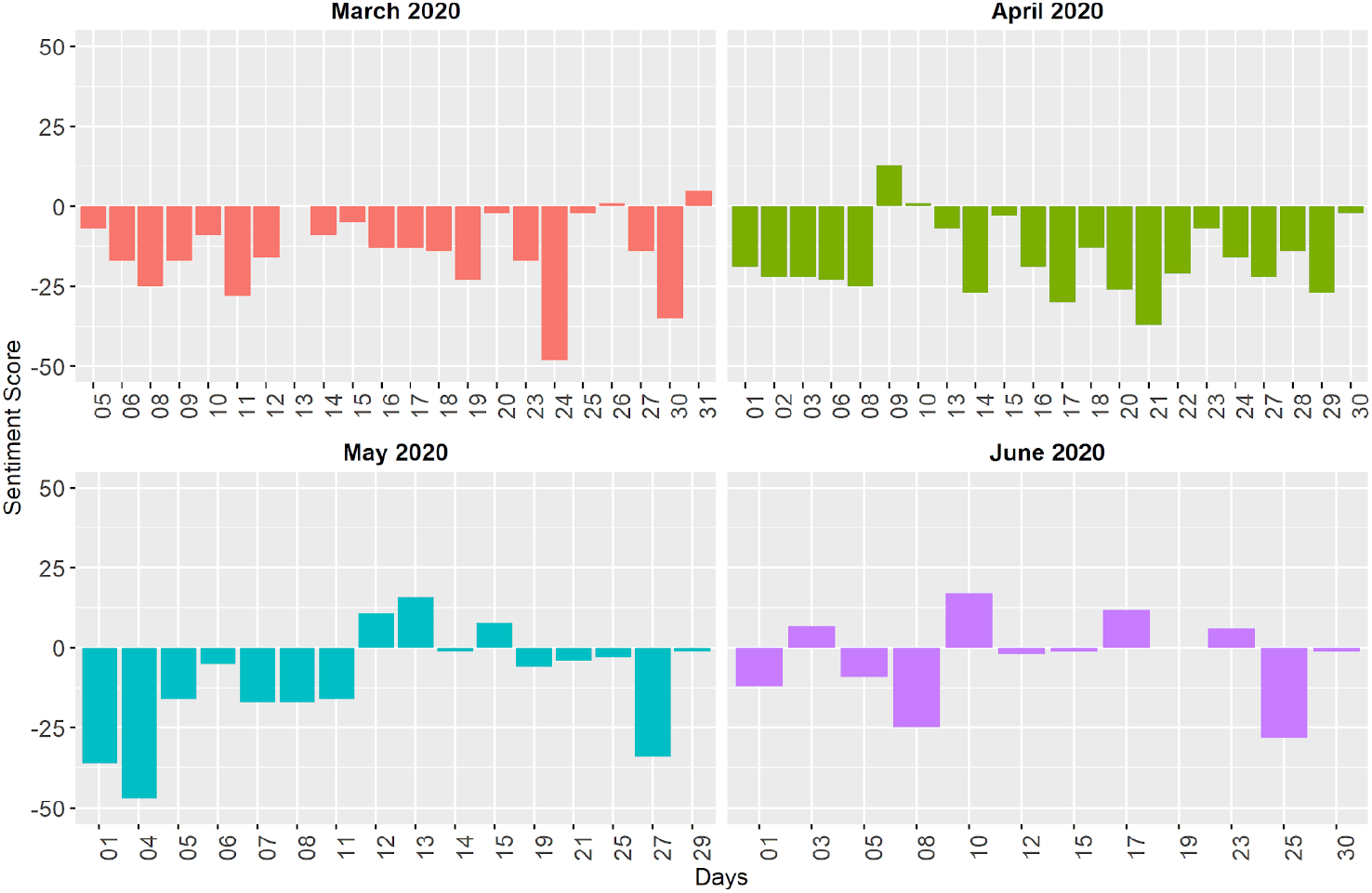
Distribution of Sentiment Scores in the Media Briefings Across the Months. *Note*. The rectangles define the time periods related to the key fluctuations and the zero line defines negative (below) and positive (above) sentiment.

**Figure 3.**
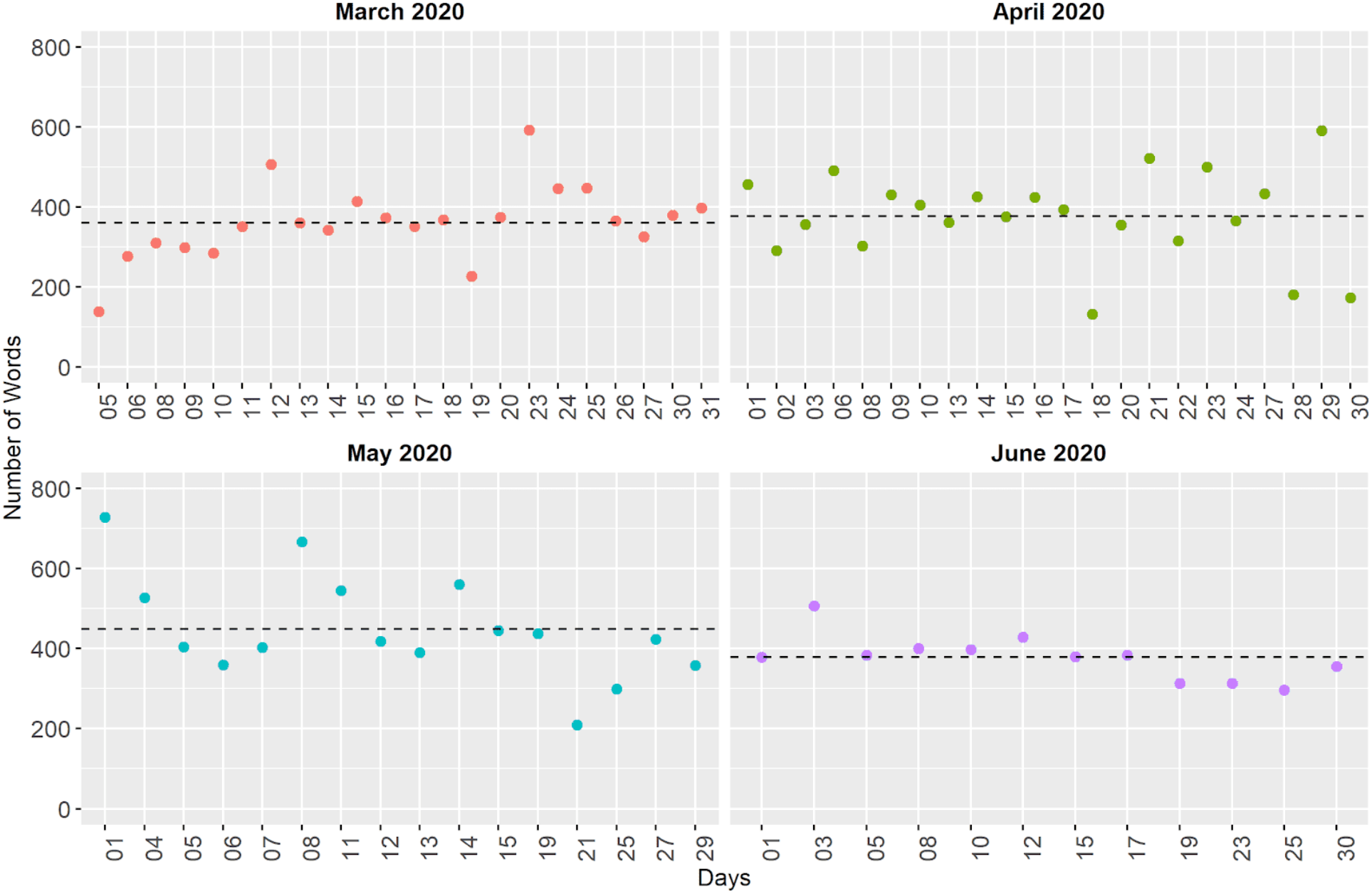
Word Counts in the Media Briefings Across the Months. *Note*. The rectangles define the time periods related to the key fluctuations and the dashed line represents the average number of words per month.

### Phase 2: Case Description from Cross Analysis of Integrated Findings

The complex case description begins with a general orientation to the pandemic response in Canada, presents the three meta-inferences derived from the cross analysis of the integrated findings at six key fluctuation points (see Table 3), and concludes with a brief update of the pandemic response since the study concluded.

**Table 3.**
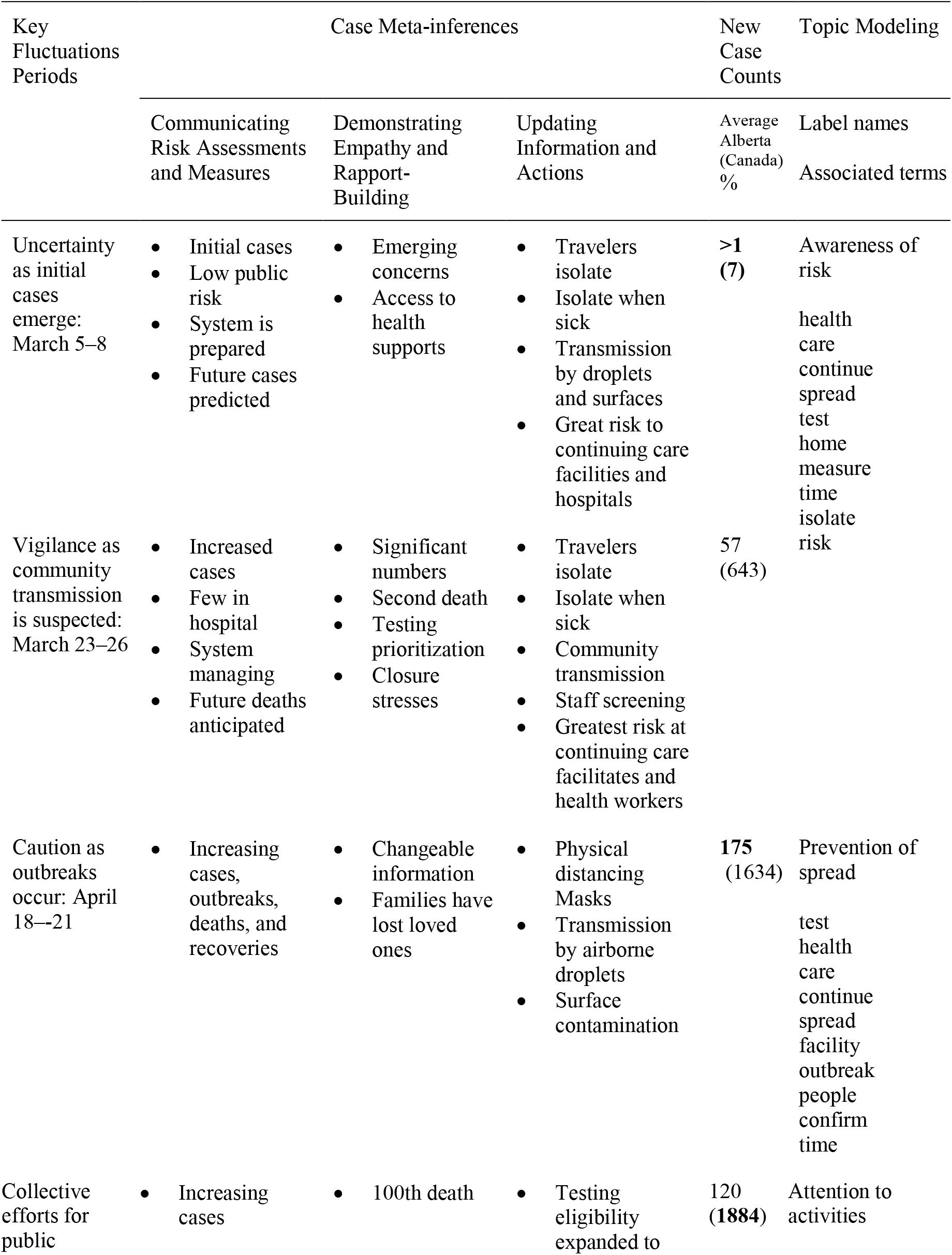
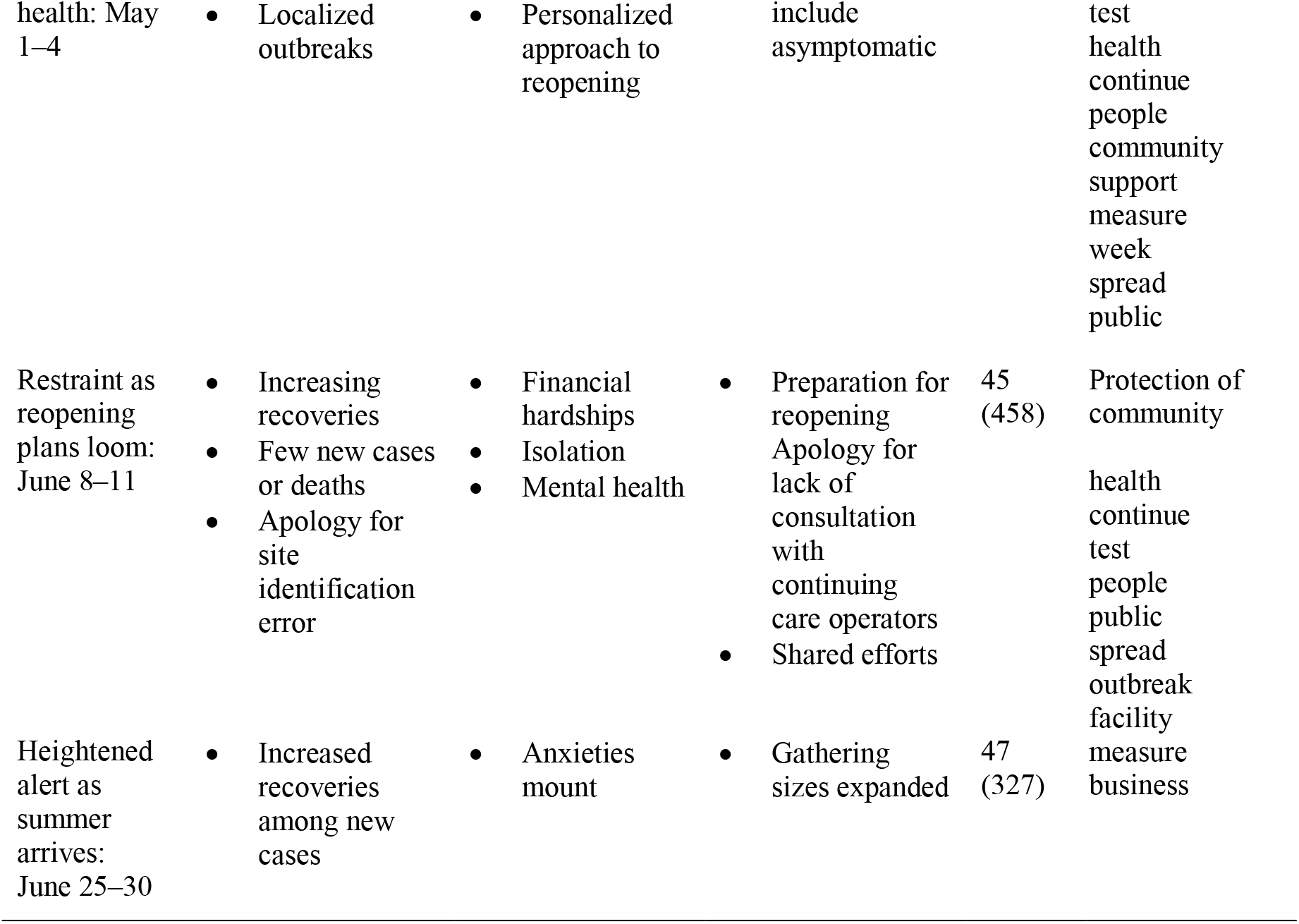
Joint Display Summary of Cross Analysis of Integrated Findings at Key Fluctuation Periods.

| Key Fluctuations Periods | Case Meta-inferences |  |  | New Case Counts | Topic Modeling |
| --- | --- | --- | --- | --- | --- |
|  | Communicating Risk Assessments and Measures | Demonstrating Empathy and Rapport-Building | Updating Information and Actions | Average Alberta (Canada) % | Label names<br>Associated terms |
| Uncertainty as initial cases emerge: March 5–8 | Initial cases<br>Low public risk<br>System is prepared<br>Future cases predicted | Emerging concerns<br>Access to health supports | Travelers isolate<br>Isolate when sick<br>Transmission by droplets and surfaces<br>Great risk to continuing care facilities and hospitals | **>1 (7)** | Awareness of risk<br>health care continue spread test home measure time isolate risk |
| Vigilance as community transmission is suspected: March 23–26 | Increased cases<br>Few in hospital<br>System managing<br>Future deaths anticipated | Significant numbers<br>Second death<br>Testing prioritization<br>Closure stresses | Travelers isolate<br>Isolate when sick<br>Community transmission<br>Staff screening<br>Greatest risk at continuing care facilitates and health workers | 57 (643) |  |
| Caution as outbreaks occur: April 18–21 | Increasing cases, outbreaks, deaths, and recoveries | Changeable information<br>Families have lost loved ones | Physical distancing<br>Masks<br>Transmission by airborne droplets<br>Surface contamination | **175 (1634)** | Prevention of spread<br>test health care continue spread facility outbreak people confirm time |
| Collective efforts for public | Increasing cases | 100th death | Testing eligibility expanded to | 120 (1884) | Attention to activities |
| health: May 1–4 | Localized outbreaks | Personalized approach to reopening | include asymptomatic |  | test health continue people community support measure week spread public |
| Restraint as reopening plans loom: June 8–11 | Increasing recoveries<br>Few new cases or deaths<br>Apology for site identification error | Financial hardships<br>Isolation<br>Mental health | Preparation for reopening<br>Apology for lack of consultation with continuing care operators<br>Shared efforts | 45 (458) | Protection of community health continue test people public spread outbreak facility measure business |
| Heightened alert as summer arrives: June 25–30 | Increased recoveries among new cases | Anxieties mount | Gathering sizes expanded | 47 (327) |  |

### The Pandemic Leadership Response Prior to March 5

The context changed rapidly during the first three months of 2020. The risk assessment for Canadians remained low on January 3, when Dr. Tam reported no cases of COVID-19 in Canada and noted: “It is important to take this seriously, and be vigilant and be prepared. But I don’t think there’s reason for us to panic or be overly concerned,” (Slaughter, 2020). By the end of January, the WHO had declared a global health emergency and Canada’s first three cases had been reported along with further reassurances from Canada’s Prime Minister, Justin Trudeau, of the measured responses the government was taking: “I can reassure Canadians that the health risk to Canadians continues to be low. We are taking all necessary precautions to prevent the spread of infection… Preventative measures are in place in airports” (Staples, 2020c). By mid-February, Dr. Hinshaw, on February 14—a month before the Canadian border would close and weeks before the first case would be detected in Alberta— communicated information about the virus, provided directions about needed actions, and called for unity in our response, stating: “Whatever the future of coronavirus, we are stronger together. Don’t let the virus divide us” (Staples, 2020b). On March 5, Dr. Deena Hinshaw began providing regular public health briefings at 3:30 in the afternoons. She assured Albertans of their preparation, saying during the initial briefing: “we have been preparing for this since the first virus emerged in January.”

### Case Meta-inference 1: Communicating Risk Assessments and Measures

Dr. Hinshaw began each briefing by updating the numbers of new cases, total cases, hospitalizations, and deaths. Figure 4 compares the low consistent rates of new case trends in Alberta with the greater fluctuation in total new cases in Canada. During the initial briefings and throughout the month of March, Dr. Hinshaw repeatedly cautioned to isolate following travel and when not feeling well. Dr. Hinshaw led by example when, in mid-March, she, herself came down with a cold, which was not COVID-19 related, and she described the situation to model how to self-isolate properly (Staples, 2020b). She made a point about each person’s contribution to manage risks as best as possible: “I encourage every Albertan to take an active role in their health and our collective health as we respond to this pandemic.”

**Figure 4.**
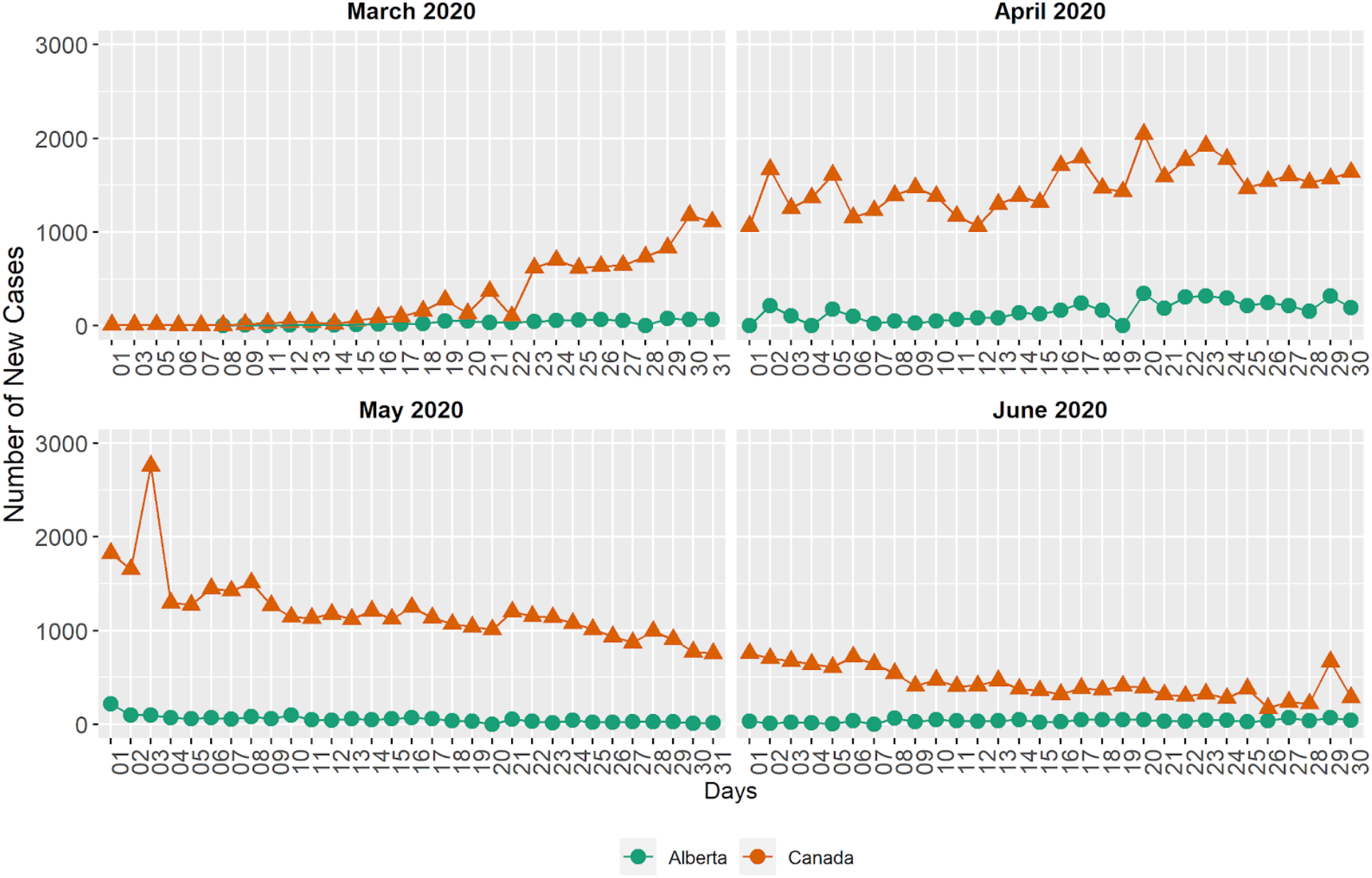
Comparison of New Daily Confirmed COVID-19 Case Trends Between Alberta and Canada. *Note*. The rectangles define the periods related to the key fluctuations, the triangles indicate total Canada data, and Alberta’s data is marked by dots.

As cases initially increased in Alberta, the overall sentiment messaging was more negative and Dr. Hinshaw’s messaging shifted to emphasize the realities of the trends: “These are significant case numbers, and they underscore the seriousness of the situation that we face.” The lag between exposure and symptoms was seen as a threat to others: “many of the new cases that are currently being diagnosed were exposed up to two weeks ago before control measures were fully in place.” A further foreshadowing of further cases in Alberta in the coming weeks because of the evolving national and global contexts conveyed with the statement: “As you know, multiple cases of COVID-19 have been confirmed across the world including a growing number of cases here in Canada.” As the outbreaks became more numerous throughout March, the sentiment was more negative than positive and word counts increased until she announced plans to move the outbreak updates to online postings and to only focus the briefings on unusual outbreaks. Dr. Hinshaw took great care to provide accurate information evidenced by beginning one of the briefings with an apology addressing incorrect identification of an outbreak site as active when it was resolved.

Those at greatest risk were identified as elderly and residents in continuing care facilities; yet outbreaks at a social gathering eventually linked to cases across the province. This heightened the risk assessment and a shift in messaging to convey that anyone was at risk: “We continue to confirm new cases in every zone, and in all age groups. We are all being impacted by this virus.” This message was relayed around the time that Prime Minister Justin Trudeau began self-isolating on March 12, as his wife had tested positive for the virus following travel to the UK (Staples, 2020b). In providing her risk assessments, Dr. Hinshaw provided sufficient information to establish trust with her audience evidenced by the following description: “She’s [Dr. Hinshaw has] quickly become a trusted face for Albertans, calmly delivering the facts as cases of COVID-19 are confirmed in the province” (Ramsay, 2020).

### Case Meta-inference 2: Demonstrating Empathy and Rapport-Building

Dr. Hinshaw began several initial briefings with the same format; she introduced herself, identified her role, provided reassurances, and described the risk to Albertans as low. Dr. Hinshaw acknowledged deaths, anxieties, and supports both frankly and with compassion evidenced by her provision of a realistic assessment of future deaths following the first death in the province on March 19, saying: “Though we are doing everything we can to limit cases of critical illnesses and death from this serious virus, tragically we know that deaths will occur.” By late March, Hinshaw had gained national attention for her public health leadership and was described as having “become a sort of guardian angel figure. She gives daily briefings, which are broadcast live and in which she manages to sound calm, kind and compassionate while being completely open and truthful about the number of people infected and the precautions that must be taken to prevent further spread” (Steward, 2020).

Throughout the case, Dr. Hinshaw described sources of anxiety as stemming from the constantly changing testing guidelines and the influx of information: “Many may feel overwhelmed and unsure of what’s important to know and what information has changed.” Dr. Hinshaw’s timing of addressing the general strain being experienced by families by the school and non-essential business closures in mid-March closely aligned with the news reporting and Alberta’s relaunch plans (Johnson, 2020; Government of Alberta, 2020b). In her announcing of reopening Dr. Hinshaw recognized that individuals may be experiencing different reactions: “We have all been affected by this pandemic. It’s completely natural to feel overwhelmed, sad or scared. You are not alone in feeling this way.” Dr. Hinshaw repeated information about mental health supports several times and provided a range of specific ideas for managing the isolation in the briefings ranging from getting outside (while maintaining distance), using video chats, and a new concept of ‘cohort families’ where two families agree to isolate from others and to focus on supporting one another: “Many Albertans out there feel alone, are afraid, or are unable to go out because they are at high risk of infection. I encourage all of us to reach out to members of the community who may be in this position.” By openly referencing both general and more specific discomforts, Dr. Hinshaw connected with the public in ways that were described by others as: “…[Hinshaw] has always shown calmness, compassion and leadership that mark her daily COVID-19 updates… Hinshaw has been widely commended for her calm and measured delivery in press briefings, her solid command of the pandemic response and genuine expressions of empathy for those suffering from the disease” (McMaster, 2020).

### Case Meta-inference 3: Updating Information and Actions

Dr. Hinshaw made a declaration that she would keep to keep the public informed: “We will also continue to be transparent and keep Albertans fully informed of any developments.” Dr. Hinshaw took care to take responsibility of any missteps evidenced by an apology addressing a lack of consultation with and notification to operators about changes to visitor policy at continuing care facilities. Areas of developments in new understandings involved that transmission was possible when asymptomatic, testing was critical for tracing and isolation, and preventive measures including enhanced screening.

Canadians were advised to stay home and avoid all non-essential travel outside of Canada in mid-March (Staples, 2020c) and around the world borders were closing including the U.S.-Canada border to non-essential travel on March 25 (Staples, 2020d). The Canadian government passed emergency legislation and introduced the Canada Emergency Response Benefit (known as CERB) (Government of Canada, 2020c) and invoked the Quarantine Act on March 26 which mandated all returning travellers to isolate for 14 days on March 26 (Staples, 2020d). Strict measures for social distancing and mass gatherings were introduced in March and described in Alberta’s Pandemic Response document (Bench, 2020). The guidelines to distance, wash hands, stay home when sick, and cover coughs and sneezes remained constant and aligned with news sources. Other preventive measures evolved over time, in response to cases—such as restricting the size of outdoor gatherings in response to case numbers: “We are making this shift based on several factors, including an analysis of how the virus is spreading in Alberta and the reduced risk of gatherings held outdoors.”

The associated words generated by the topic modelling suggest a shift in messaging about emerging understandings: in March the focus was on awareness of risk, and by June, it shifted to protection of community. It is also interesting to note that half of the associated words are common to those two months (see Table 3). Many of the unique words refer to the prevention measures in place at the time; for example, in March the messages conveyed the need to stay *home, isolate* and take the *time* to *care* for those at *risk* (associated words are italicized) whereas in June the messages expressed the need to protect the p*ublic, people, businesses*, and *facilities* from *outbreaks*.

Dr. Hinshaw continually highlighted contributions of the government; their planning and mobilization efforts early on, expanding testing access, and launching the mobile-based tracing technology in May. There were clear connections between the comments about needing to work together to prevent the spread of infection and the associated words generated by the topic modelling. This suggests consistency messaging about needed actions and cautions in both April and May as the common words related to *continue* to *test*, avoid *spread* and *care* for *people* (associated words are italicized); whereas the unique words in April reflected what was going on at the time related to outbreaks at facilities, and then in May the focus shifted to supporting community. As well Dr. Hinshaw regularly recognized the efforts of Albertans as a whole and groups of individuals for their contributions to the public health response: “Every act–those big or small–lifts us up as a province and demonstrates how quickly Albertans take action to care for one another,” and “I want to end today by saying how grateful I am to everyone for following the public health guidelines…. Let’s continue to take pride in that protection of each other as we move forward—to enjoy our activities, but with caution and care for those around us.” Dr. Hinshaw’s masterful ability to point out how the actions can benefit or protect the larger community is evidenced by the description bestowed upon her by others, “We now count on Hinshaw. Always reassuring with her quiet, steady delivery. Always reminding us of the simple steps to protect ourselves. Always urging us to do the right thing, though never scolding” (Staples, 2020b).

### The Ongoing Pandemic Response Since July 5

On July 5, the number of new cases had stabilized and Dr. Deena Hinshaw stopped providing regular public health briefings. At the time of this writing (September 22, 2020), the pandemic is ongoing and it is yet unknown when the public health crisis will cease to require a coordinated response. As the weather turns colder as winter approaches, there are increased concerns in Alberta about indoor exposure risks.

## Discussion and Implications

### Contribution to Public Health Communication Literature

The complex case description reveals novel insights about how public health briefings can build credibility and trust within the rapidly evolving context of a global pandemic to address the phase 2 guiding research question (see Table 1). Specifically, we see the masterful weaving of three key functions of a chief public health officer related to risk assessments and communications, demonstrations of empathy, and actionable information updates (see Table 4).

**Table 4.**
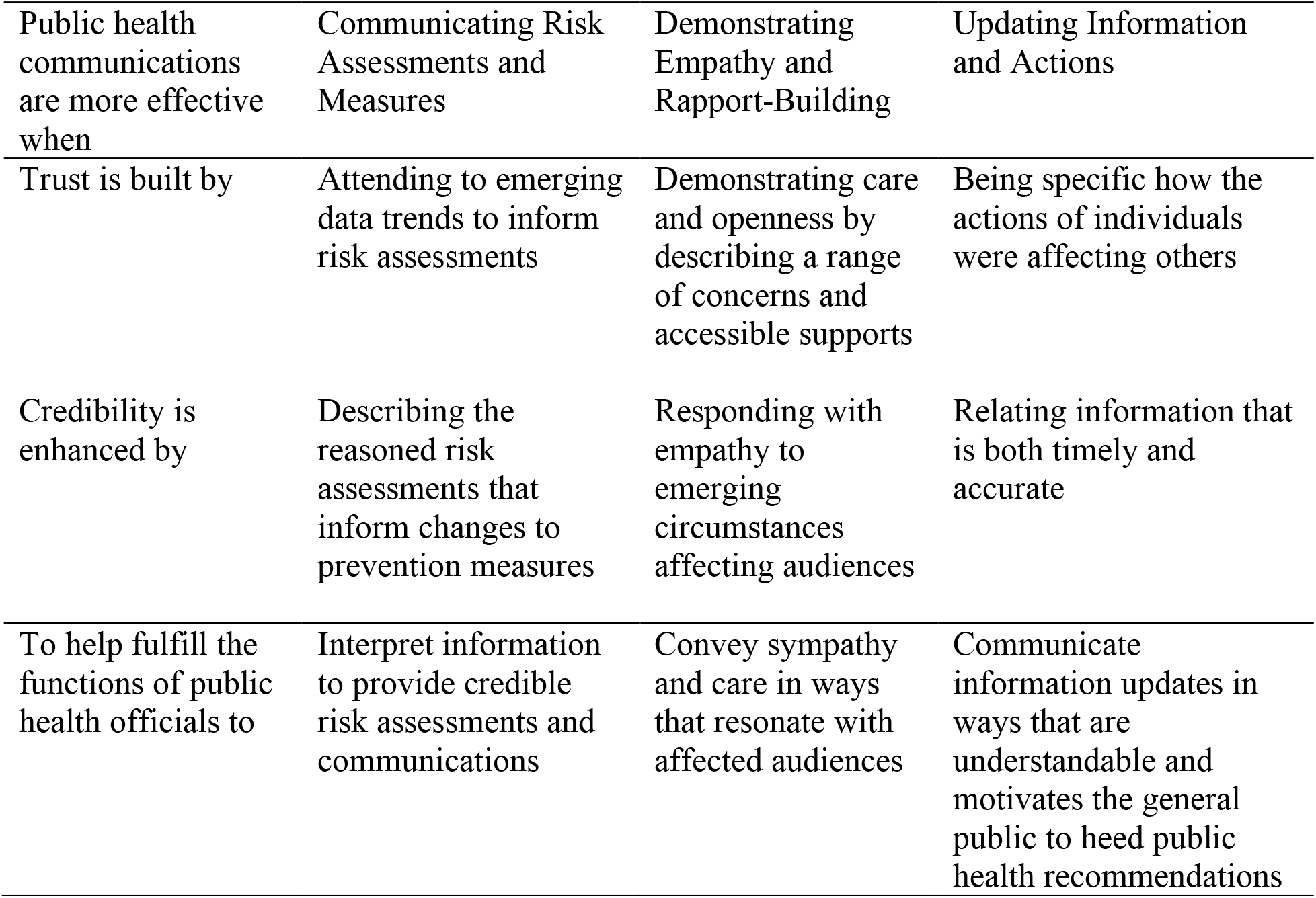
Contributions to Public Health Communications Literature.

The novelty of the virus and the challenges inherent to predicting transmission rates meant that interpreting case data trends was an essential part of the chief public health officer’s function to provide credible risk assessments and communications (CDC, 2018). Our case description provides evidence of the consistent use of the same case statistics from a trusted institution to inform shifts in risk assessments and the provision of reasoned interpretations of the data as informing changes to the recommended prevention measures. The risk assessments and communications were effective when the changes to prevention measures were accompanied by detailed explanations that the shifts to risk assessments were responding to emerging data trends from trusted sources.

The unprecedented impacts on the daily interactions of the public meant that explicitly recognizing the far-reaching, changeable, and unique effects on individuals and groups was a key feature of the chief public health officer’s function to demonstrate empathy in ways that resonate with affected audiences (Tumpey et al., 2018). Our case description offers many illustrative examples of both shared and unique impacts of the virus; from new concerns for loved ones living in long term care facilities and stresses for families with school closures to new discomforts associated with physical distancing measures and financial anxieties arising from business closures. The demonstrations of empathy through expressing sympathy and describing supports became more candid and specific over time as rapport with the Alberta public was built.

Updating the public on the rapidly changing information surrounding the virus meant communicating in ways that were understandable and timely was a necessary focus of the chief public health officer’s work in motivating the general public to heed public health recommendations (CDC, 2018; WHO, 2012). Our case description draws attention to the essential role for the briefings to be specific in how the actions affected others and timely as well as aligned with other information sources. The information was more persuasive when the benefits of individual and collective actions to protect the larger community was communicated explicitly and reflected what was also being reported in the news.

### Contributions to Mixed Methods Research Practice

This study contributes an illustrative example and discussion for guiding how a mixed methods convergent sequential research design, informed by complexity theory and drawing upon open-access datasets, can rapidly generate complex case study descriptions. Specifically, the article advances mixed methods research practice in two ways and in so doing heeds the calls for illustrative examples of novel combinations of data procedures constituting methodological advancements (Creswell & Plano Clark, 2018; Mertens et al., 2016). First, the use of text mining to manage large volumes of open-access data efficiently for the purpose of identifying atypical periods of sentiment fluctuation to serve as the points of integration within a sequential convergent mixed methods design. Our generation of a holistic complex case description from the cross analysis of the integrated findings at the six key fluctuation periods mitigated the limitations of either qualitative or quantitative approaches alone more efficiently than had we collected the data ourselves or analyzed the entire case dataset. Second, the use of a complexity-informed CS-MM design to study complex phenomena for the purpose of generating novel public health insights aligns with one of the four theoretical conceptions of complexity theory for mixed methods researchers advanced by Kallenmeyn and colleagues (2020). Our CS-MM design procedures, informed by complexity theory, provide practical guidance for documenting the emergent, interdependent, and adaptive realities of the initial public health response to the COVID-19 pandemic. Together, these collective insights along with drawing upon the interdisciplinary and diverse methodological expertise of the researchers involved in the current study, we provide an essential reference for others to learn from and build upon.

### Strengths, Limitations, and Future Directions

Reliance on open access data creates new opportunities for the use of text mining techniques within mixed methods research designs as an efficient means to detect trends within large datasets. While the use of media briefings is unique as a means of accessing the message conveyed by public health authorities, it is also limited because we do not have data about the impacts of the messages on the public’s behaviour. Future studies could include interviews with the public health authorities involved in the creation and delivery to seek an ‘insider’ perspective, including Dr. Deena Hinshaw herself, if possible. Our case boundary of the onset period of the pandemic should be considered in light of the fact the pandemic is ongoing at the time of writing, and global public health response and communication efforts are continuing. Future studies could expand the case boundaries to include further data sources and time periods.

## Conclusion

The integration of publicly accessible data during the unprecedented and rapidly changing situation surrounding the onset of the COVID-19 pandemic in Alberta (Canada) contributes to novel insights about how public health briefings can build credibility and trust within the rapidly evolving context of a global pandemic. Specifically, we provide practical examples relating three key functions of a chief public health officer related to risk assessments and communications, demonstrations of empathy, and actionable information updates for enhancing the effectiveness of public health communications during the onset of a pandemic. The current article demonstrates how text mining procedures within a mixed methods convergent sequential research design, informed by complexity theory and drawing upon open-access datasets, rapidly generate complex case study descriptions. This study highlights the usefulness of conceptualizing the public health response to the COVID-19 pandemic as a complex adaptive system and calls for further application of complexity science in mixed methods research designs.

## Data Availability

Public health communications hosted by Dr Deena Hinshaw media briefing transcripts were downloaded in Word format from the government of Alberta COVID 19 pandemic website
daily new Alberta confirmed cases of COVID 19 case statistics were downloaded and aggregated in excel spreadsheet format from the government of Alberta interactive COVID 19 data application website

https://www.alberta.ca/stats/covid-19-alberta-statistics.htm

https://www.alberta.ca/covid

## Acknowledgements

We are grateful to the Government of Alberta’s efforts for open access data. This work was supported by funding from the University of Alberta’s Endowment Fund for the Future – Support for the Advancement of Scholarship Program. We would like to thank Adrienne Montgomery for helpful comments during the review of a draft of this manuscript.

## Author Biographies

**Cheryl N. Poth** (Ph.D., Queen’s University, Kingston, Ontario, Canada) is an applied education and health research methodologist and a professor of Measurement, Evaluation, and Data Science at the University of Alberta. She is also a member of the Centre for Research in Applied Measurement and Evaluation (CRAME) in the Faculty of Education and adjunct professor in the Faculty of Medicine and Dentistry at the University of Alberta. Her research interests, scholarship, and teaching include enhancing teaching and learning of mixed methods and qualitative research and generating complexity-informed methodological innovations.

**Okan Bulut** (Ph.D., University of Minnesota, Minneapolis, Minnesota, USA) is an Associate Professor of Measurement, Evaluation, and Data Science at the University of Alberta. He is also a member of the Centre for Research in Applied Measurement and Evaluation (CRAME) in the Faculty of Education at the University of Alberta. Dr. Bulut’s research interests include survey design, educational data mining, and statistical data analysis and visualization using software programs like R.

**Alexandra M. Aquilina** (M.Ed., University of Alberta, Edmonton, Alberta, Canada) is a doctoral student in the School and Clinical Child Psychology program at the University of Alberta. Her research interests include investigating the assessment of cognitive abilities, particularly in the context of culturally diverse students, and the influence of teacher beliefs about the academic potential of students with disabilities. She has a particular methodological interest in qualitative and mixed methods research.

**Simon J. G. Otto** (Ph.D., University of Guelph, Guelph, Ontario, Canada; D.V.M, University of Saskatchewan, Saskatoon, Canada) is veterinary epidemiologist and an Associate Professor of epidemiology at the University of Alberta. His research interests focus on the One Health epidemiology of AMR, including surveillance and genomic diagnostic methods to inform antimicrobial stewardship policy. He has worked multiple large foodborne illness outbreak responses, conducted scientific knowledge synthesis for COVID-19 policy to advise Alberta Health Services (AHS), and sits on an AHS zone Emergency Outbreak Management Task Force for COVID-19.

